# Validity of a smartphone app to objectively monitor performance outcomes in degenerative cervical myelopathy: an observational study

**DOI:** 10.1101/2023.09.13.23294900

**Authors:** Alvaro Yanez Touzet, Tatiana Houhou, Zerina Rahic, Ilya Laufer, Konstantinos Margetis, Allan R Martin, Nicolas Dea, Zoher Ghogawala, Misha Kapushesky, Mark R N Kotter, Benjamin M. Davies, MoveMed

## Abstract

**Objectives:** To assess the validity of MoveMed, a battery of performance outcome measures performed using a mobile phone application, in the measurement of degenerative cervical myelopathy (DCM).

**Design:** Prospective observational study.

**Setting:** Decentralised secondary care in England, United Kingdom.

**Participants:** 27 adults aged 60 (SD: 11) who live with DCM and possess an approved smartphone.

**Primary and secondary outcome measures:** Criteria from the Consensus-based Standards for the selection of health Measurement Instruments (COSMIN) manual were used to assess validity and risk of bias. Briefly, each MoveMed outcome was compared to two patient-reported comparators, and a priori hypotheses of convergence/divergence were tested against consensus thresholds. The primary outcome was the correlation coefficient between the MoveMed outcome and the patient-reported comparators. The secondary outcome was the percent of correlations in correspondence with a priori hypotheses. The comparators were the patient-derived modified Japanese Orthopaedic Association (P-mJOA) score and the World Health Organization Quality of Life Brief Version (WHOQOL-Bref) questionnaire. Thresholds for convergence/divergence were ≥0.3/<0.3, and >0/<0 for directionality.

**Results:** As expected, MoveMed’s tests of neuromuscular function correlated most with questionnaires of neuromuscular function (≥0.3) and least with questionnaires of quality of life (<0.3). Furthermore, directly related constructs correlated positively to each other (>0), while inversely related constructs correlated negatively (<0). Over 70% and 50% of correlations (unidimensional and multidimensional, respectively) were in accordance with hypotheses. No risk of bias factors from the COSMIN Risk of Bias checklist were recorded. Overall, this was equivalent to ‘very good’ quality evidence of sufficient construct validity in DCM.

**Conclusions:** Criteria from COSMIN provide ‘very good’ quality evidence of the validity of the MoveMed tests in an adult population living with DCM.

## Strengths and limitations of this study

□ Criteria from the COSMIN manual were used to assess the validity of the MoveMed battery of performance outcome measures.
□ Criteria from the COSMIN manual rate the methodological quality of the evidence as ‘very good’.
□ Self-reported outcome measures and data elements were used in a decentralised secondary setting.
□ A single primary outcome was used to assess validity.
□ Study design and analyses were performed by individuals formally trained in clinimetrics.

## INTRODUCTION

Abnormal limb movement is a key phenotype of disease affecting the nervous and musculoskeletal systems. Loss of dexterity, for example, is a notable manifestation of conditions like Parkinson’s disease, degenerative cervical myelopathy (DCM), peripheral neuropathy, and osteoarthritis (1, 2). The significance of this phenotype can be seen in the physician’s approach to examining the neuromuscular systems, the features used to distinguish or measure its disease, or the information sought to define its care and research. Collectively, diseases affecting the nervous and musculoskeletal systems are estimated to account for 1.1m to 4.9m deaths and 165m to 357m DALYs worldwide and are the leading causes of global disability, reflecting their often-chronic nature (3–5).

Whilst abnormal movement is a key component of diagnosis, it is also key a component of longitudinal monitoring, as these diseases typically lack responsive serological or imaging biomarkers. Such monitoring is key to adjust or review treatment strategies over time, and to define the success or failure of research trials (6). Today, monitoring relies on qualitative outcome measures: classifications based on a hierarchy of exemplar functions, such as questionnaires and/or item selection. Whilst qualitative tools can be robust, valid, and even performed by the patient remotely, their limited granularity and intrinsic subjectivity means they lack accurate and responsive discrimination of small but significant changes, particular for fluctuating disease (7). For clinical care, this means clinically important change is seen late, often at the cost of increased disability. For clinical research, the low statistical power of qualitative tools means far higher sample sizes are needed for trials to mitigate Type 2 error.

This is exemplified within DCM, a slow-motion spinal cord injury estimated to affect 1 in 50 adults (8–11). Here, dexterity, gait, and balance are key measurement constructs (12). Currently, the gold-standard outcome measure is the modified Japanese Orthopaedic Association Score (mJOA), but it is poorly responsive (13). Further, score variation, driven partly by the disease, and partly by reliability, is more than twice the minimal clinical important difference. In practice, this demands sample sizes greater than 300 patients for 1:1 comparison with at least 80% power (14, 15). Developing new approaches to functional measurement is a recognised research priority (12).

Advances in our ability to assess limb performance can thus greatly improve our understanding of the patient’s clinical picture, lead to better decision-making and outcomes, as well as accelerate knowledge discovery (16, 17). The sensors contained within smartphones offer the potential to achieve this. Smartphones are increasingly carried by all patient groups, with far greater penetrance and priority than other wearable devices such as smartwatches (18). Current focus in portable technology with respect to health has largely been on ‘background monitoring’, but shortcomings remain, including accurate and responsive insights at the individual patient level, as well as between-device variation (19).

This study evaluates MoveMed, a smartphone application originally developed by researchers from the University of Cambridge to assess hand, arm, and leg function in real-time, in the user’s natural environment, and under standardised conditions. This approach is therefore different from background monitoring: it harnesses the accuracy of mobile sensors to measure movement but does so during prescribed activities or tasks, designed by healthcare professionals and patients to target critical markers of disease. It can therefore be considered a patient-performed, performance outcome measure (PerfO or PerfOM). Since MoveMed is being developed in accordance with ISO 13485 (Software as a Medical Device), testing of measurement properties was timely given recent laboratory experience of technological readiness (TRL4). In terms of V3 stages for biometric monitoring technologies (BioMeTs) (20), the testing in this article corresponds to clinical validation.

MoveMed was originally developed for DCM. Therefore, the focus of this report is on the validity of the MoveMed battery of PerfOM’s in DCM. However, recognising that the measurement constructs in this disease are shared across other neuromuscular disease, its validity is currently being explored in other conditions. Formal methods and criteria from the United States Food and Drug Administration (FDA) and Consensus-based Standards for the selection of health Measurement Instruments (COSMIN) guidelines were used to design a prospective and decentralised observational study. Validity and risk of bias were principally assessed via hypothesis testing of construct validity. Content validity will be formally evaluated separately but is briefly described in this work. This article is the first of a series of clinimetric studies about the measurement properties of MoveMed’s battery of PerfOM’s.

## METHODS AND ANALYSIS

### Participants

Between September 2022 and April 2023, 27 people living with DCM were enrolled in the prospective and decentralised EMPOWER study (www.movemed.io/empower). Prospective participants were recruited via an online campaign and asked to complete consent and registration forms (Figure 1) (21, 22). These were used to screen participants for eligibility. Participants were deemed eligible if they had a self-reported diagnosis of DCM, owned a smartphone, and were able to stand and walk without the assistance of another person. Eligible participants were invited to download the MoveMed App to their smartphones and complete an electronic, baseline questionnaire on neuromuscular function, hand dominance, and quality of life. This included questions from the patient-derived mJOA (P-mJOA) and the World Health Organization Quality of Life Brief Version (WHOQOL-Bref).

**Figure 1.**
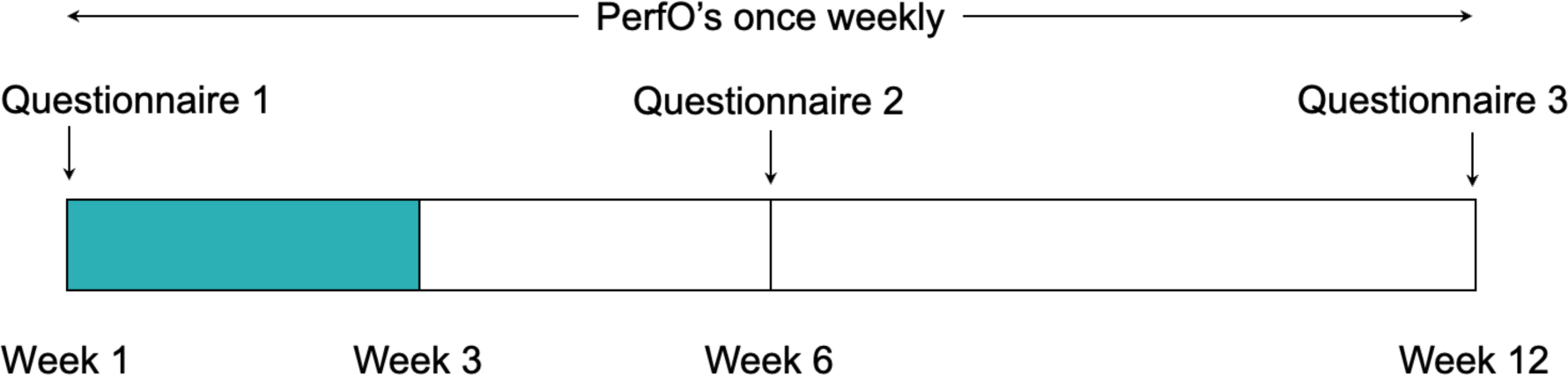
Study timeline. Data from the first three weeks of the study were included due to the on-going status of the trial.

All enrolled participants were asked to perform each task in the MoveMed App once per week for a period of 12 weeks. Task adherence was remotely monitored once a week using a bespoke online dashboard. Participants were offered reminders and help via email once a week if 14 days passed since the completion of the latest task. These were offered for a total of two consecutive times per participant, after which the participant was considered lost to follow-up. At weeks six and 12, participants were asked to complete the same electronic questionnaire from week one.

### MoveMed and tasks

MoveMed is a smartphone application designed by academic neurosurgeons and computer scientists from the University of Cambridge to administer PerfOM’s (Figure 2). These may be administered by clinicians during in-person visits or self-performed by individuals in community. Version 1.0.0 of the App originally offered three performance tasks: a Fast Tap Test, a Hold Test, and a Stand and Walk Test. Version 1.2.2 incorporated an additional offering—a Typing Test—while making no changes to the three original tasks. Versions 1.0.0 and 1.2.2 were respectively available in the Android Google Play Store and iOS App Store at the time of writing and were used in this study by enrolled participants.

**Figure 2.**
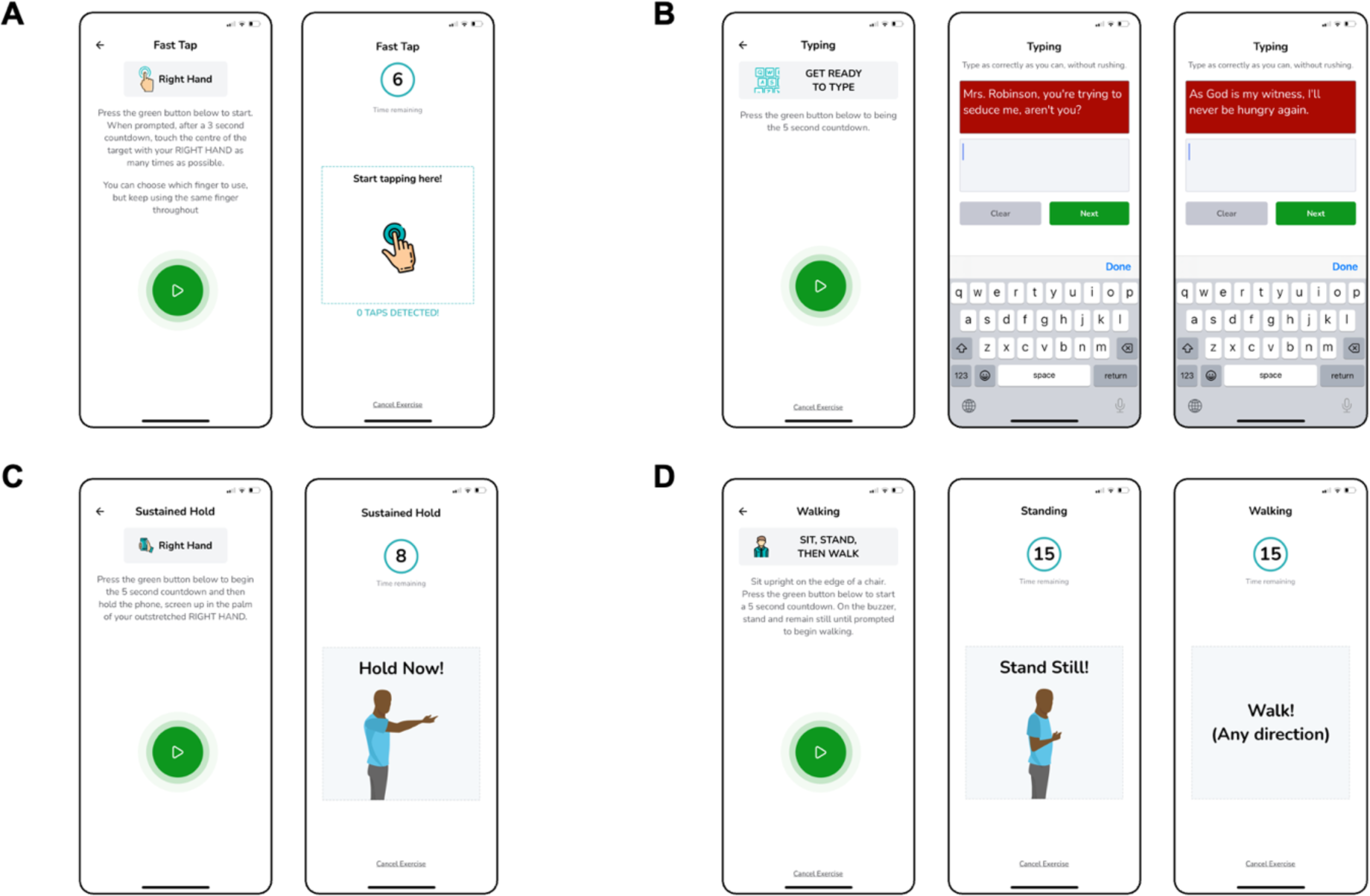
Schematic illustrations of the MoveMed battery of performance outcome measures. A: The six-second fast-tap test. B: The two-stage typing test. C: The eight-second sustain-hold test. D: The 15-second stand and walk test.

The Fast Tap Test is a unidimensional PerfO task that assesses finger dexterity through an interactive, six-second smartphone task. Users are shown a demonstrative cartoon (Figure 2A) and instructed to ‘touch the centre of the target with [each] hand as many times as possible’. In-app video demonstration is also available. The construct (finger dexterity) is assessed by measuring the speed, accuracy, and efficiency of finger tapping as continuous variables and analysing them as a panel of unidimensional measures. Content validity was assessed by AYT, MRNK, and BMD through literature review and clinical and patient input and deemed relevant, comprehensive, and comprehensible at the time of development (23–25). In this study, tap latency was used as a reflective measure of finger dexterity.

The Typing Test is another unidimensional PerfO task that assesses finger dexterity through an interactive, two-stage smartphone task. Users are shown a demonstrative cartoon (Figure 2B) and instructed to ‘type as correctly as they can, without rushing’. In-app video demonstration is also available. The construct (finger dexterity) is assessed by measuring speed, accuracy, and efficiency of typing as continuous variables and analysing them as a panel of unidimensional measures. Content validity was assessed by AYT, MRNK, and BMD through literature review and clinical and patient input and deemed relevant, comprehensive, and comprehensible at the time of development (23–25). In this study, typing speed was used as a reflective measure of finger dexterity.

The Hold Test is a unidimensional PerfO task that assesses upper limb stability through an interactive, eight-second smartphone task. Users are shown a demonstrative cartoon (Figure 2C) and instructed to ‘hold the phone, screen up in the palm of [their] outstretched hand’. In-app video demonstration is also available. The construct (upper limb stability) is assessed by measuring the involuntariness, rhythmicity, and oscillation of the upper limbs as continuous variables and analysing them as a multidimensional Stability Score. Content validity was assessed by AYT, MRNK, and BMD through literature review and clinical and patient input and deemed relevant, comprehensive, and comprehensible at the time of development (23–25). In this study, the Stability Score was used as a reflective measure of upper limb stability.

The Stand and Walk Test is a multidimensional PerfO task that assesses gait through an interactive, two-stage smartphone task. During the first stage, users are instructed to ‘sit upright on the edge of a chair [and to] press the green button [when they are ready to] stand and remain still’. During the second stage, users are instructed to ‘walk [in] any direction’. In-app cartoon and video demonstration are also available (Figure 2D). The construct (gait) may then be assessed by measuring standing or walking as continuous variables and analysing them as multidimensional measures. Content validity was assessed by AYT, MRNK, and BMD through literature review and clinical and patient input and deemed relevant, comprehensive, and comprehensible at the time of development (23–25). In this study, cadence was used as a reflective measure of gait.

### Patient-reported comparators

Two patient-reported outcome measures (PRO’s or PROM’s) were used as comparators for DCM: the P-mJOA and the WHOQOL-Bref.

The P-mJOA score is a multidimensional, patient-reported questionnaire that assesses neuromuscular function in DCM across four items: motor function of the upper and lower extremities (MDUE and MDLE, respectively), sensory function of the upper extremities, and sphincter function (26). Responses are scored on an ordinal scale per item and presented as both a panel of unidimensional scores and an unweighted sum-total, multidimensional score. The P-mJOA was selected due to the existence of systematic assessment of construct validity (*r* > 0.5) and feasibility in DCM (13) and due to the use of its clinically reported analogue (the mJOA) as the current gold standard. The P-mJOA was favoured over the mJOA since it is intended to be a truly patient-reported equivalent of the mJOA, which can be understood by individuals with no medical knowledge or training (27).

The WHOQOL-Bref is a multidimensional, patient-reported questionnaire that assesses quality of life across 26 items grouped into four domains: physical health (PH), psychological health (PS), social relationships (SR), and environment health (EH) (28). Responses are scored on a five-point ordinal scale per item and presented as a panel of sum-total, multidimensional scores. Responses to two items may, furthermore, be presented individually to give insight into the respondent’s global perception of their quality of life and their quality of health. These were presented in writing to describe the population’s characteristics but were not considered robust enough to warrant correlation analysis. The WHOQOL-Bref was selected due to the existence of systematic assessments of validity, reliability, and responsiveness in traumatic brain injury (29), Parkinson’s disease (30), and DCM (13). It was also favoured over the 36-Item Short Form (SF-36) Health Survey due to its relative brevity and over the EuoQOL Five Dimensions (EQ-5D) Questionnaire due to licensing restrictions.

### Statistical analysis

The COSMIN manual defines validity as ‘the degree to which a[n outcome measure] measures the construct it purports to measure’ (31). In the absence of a gold standard, validity may be assessed formally through hypotheses testing of correlations to known standards. These may then be judged both as a panel of stand-alone ratings (32).

In this study, we assessed validity by correlating the MoveMed’s PerfO’s to their corresponding patient-reported comparators. This was achieved by comparing to the P-mJOA and WHOQOL-Bref PROM’s. Due to the on-going status of the trial, data from the first three weeks of the study were included (Figure 1). All available tests within this period were included. Longitudinal replicates of MoveMed tasks were averaged before comparing their mean scores to the mean scores of the PRO’s. Responses from the baseline questionnaire were used and results were sub-grouped by diagnosis. Missing data were not imputed, and all analyses were done using Python 3.10.12.

Spearman rank correlation coefficients were computed due to their suitability for ordinal scales. In accordance with COSMIN, P values were not used because ‘the validity issue is about whether direction and magnitude of a correlation is similar to what could be expected based on the construct(s) that are being measured [and not] whether correlations statistically differ from zero’ (31, 33). Hypotheses for direction and magnitude of correlations were thus drawn separately and adapted from COSMIN (31) and de Vet et al (34). We hypothesised that:

− The magnitude of correlations between outcomes measuring similar constructs should be ≥0.5;
− The magnitude of correlations between outcomes measuring related, but dissimilar, constructs should be ≥0.3, and ideally <0.5;
− The magnitude of correlations between outcomes measuring unrelated constructs should be <0.3; and
− The direction of correlations between outcomes measuring directly related constructs should be positive (>0) and negative (<0) between outcomes measuring inversely related constructs.

As in Yanez Touzet et al (13), constructs were defined as ‘similar’ if they both measured the same domain with a unidimensional instrument. If they measured the same domain, but at least one of the instruments was multidimensional, the constructs were defined as ‘related but dissimilar’. Constructs measuring different domains were otherwise defined as ‘unrelated’.

### Risk of bias assessment

The COSMIN Risk of Bias checklist (31) was used to assess the methodological quality of hypotheses testing.

### Overall assessment

Overall assessments of construct validity were made using a panel of ratings and prior knowledge of content validity. These were appraised qualitatively and presented in writing due to the relatively higher importance of some comparators over others. As in COSMIN (31), correlations were converted into ratings by comparing results to hypotheses. Correlations in accordance with hypotheses were rated ‘sufficient’. Correlations in opposition were rated ‘insufficient’. Correlations in between boundaries (e.g., *r* = 0) and statistical artefacts (e.g., non-monotonic data) were rated ‘indeterminate’.

## RESULTS

27 participants with DCM enrolled in the prospective and decentralised EMPOWER study (Figure 3), principally via advertisement through Myelopathy.org, a DCM charity (21, 22). On average, participants were 60 years old (SD: 11; Table 1). DCM severity ranged from mild to severe (P-mJOA total score: 8–18). The impact on upper limb motor function ranged from none to ‘unable to eat with spoon but able to move hands’ (P-mJOA MDUE subscore: 2–5) and the impact on lower limb motor function ranged from none to ‘able to move legs but unable to walk’ (P-mJOA MDLE subscore: 2–7). Overall health perception ranged from ‘satisfied’ to ‘very dissatisfied’ (WHOQOL Overall health: 1–4) and overall QOL perception from ‘very good’ to ‘very poor’ (WHOQOL Overall QOL: 1–5). In terms of the MoveMed PerfO’s, participants paused for 80–2,600 ms in between taps and typed ∼0.6–2.5 keys per second. Arm stability ranged from 39% to 100% and cadence from 14 to 112 steps per minute.

**Figure 3.**
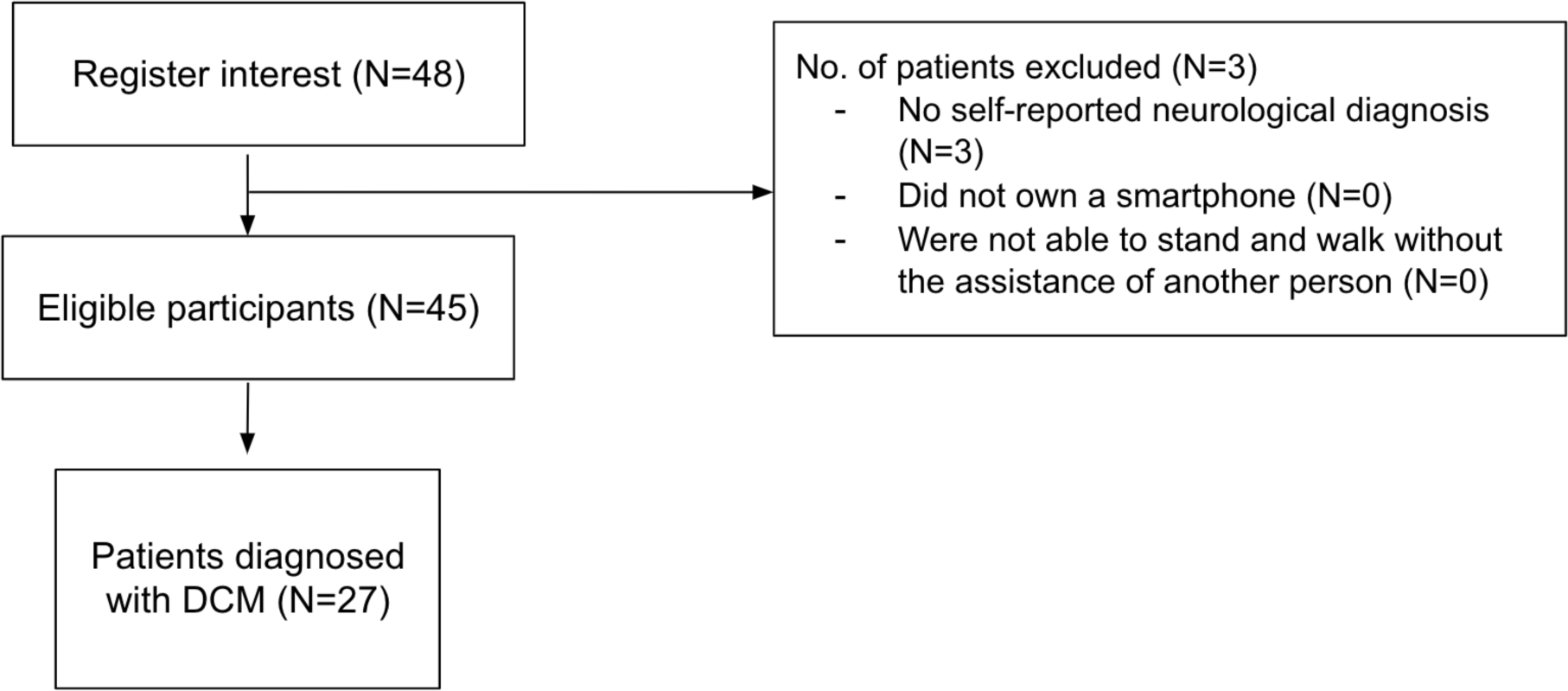
STROBE diagram.

**Table 1.** Characteristics of study participants.

| Feature | Value |
| --- | --- |
| No. of participants | 27 |
| Age | 60.7 (10.8) |
| P-mJOA (reference range: 0–18) | 11.4 (2.9) |
| MDUE (reference range: 0–5) | 3.4 (1.1) |
| MDLE (reference range: 0–7) | 4.1 (1.3) |
| SDUE (reference range: 0–3) | 1.6 (0.8) |
| SD (reference range: 0–3) | 2.3 (0.7) |
| WHOQOL-Bref | — |
| Overall QOL (reference range: 1–5) | 2.9 (1.1) |
| Overall health (reference range: 1–5) | 2.6 (0.8) |
| EH (reference range: 8–40) | 25.5 (5.5) |
| PH (reference range: 7–35) | 18.6 (5.6) |
| PS (reference range: 6–30) | 18.6 (4.1) |
| SR (reference range: 3–15) | 9.2 (2.1) |
| MoveMed Fast Tap Test Inter-Tap Duration* (s) | 0.24 (0.13), 0.36 (0.52) |
| MoveMed Hold Test Stability Score* (%) | 78.6 (16.0), 75.8 (15.1) |
| MoveMed Typing Test Speed (keys per second) | 1.39 (0.42) |
| MoveMed Stand and Walk Test Cadence (steps per min) | 58.8 (26.9) |

Data reported as *n* or mean (standard deviation) unless otherwise specified. Ranges reported in manuscript body.
\*Data reported as ‘dominant hand, non-dominant hand’ mean (standard deviation) pairs. Ranges reported in manuscript body.
EH: Environmental health, MDLE: Motor dysfunction of the lower extremity, MDUE: Motor dysfunction of the upper extremity, P-mJOA: Patient-derived modified Japanese Orthopaedic Association, PH: Physical health, PS: Psychological health, QOL: Quality of life, SD: Sphincter dysfunction, SDUE: Sensory dysfunction of the upper extremity, SR: Social relationships, WHOQOL-Bref: World Health Organization QOL Brief Version

Differential app use was noted throughout the studied period (Table 2). More participants used the Stand and Walk and Typing Tests (N≥20) than the Fast Tap and Hold Tests (N≥12). However, mean adherence was higher with the Fast Tap and Hold Tests (100% and 90%, respectively) than with the Stand and Walk and Typing Tests (77% and 72%, respectively). Crucially, median adherence was satisfactory: 100% for the Fast Tap, Hold, and Stand and Walk Tests, and 80% for the Typing Test. Differential use was thus attributed to individual test preferences.

**Table 2.** Correlations, ratings, and hypotheses for the testing of construct validity.

| MoveMed outcome measure | Comparator | Hypotheses |  | Result* | N | Rating<sup>†</sup> |  | ROB |
| --- | --- | --- | --- | --- | --- | --- | --- | --- |
|  |  | Direction | Magnitude |  |  | Direction | Magnitude |  |
| MoveMed Fast Tap Test | P-mJOA total score | $r < 0$ | $0.3 \leq \vert r\vert (< 0.5)$ | -0.47, -0.42 | 12 | +, + | +, + | No |
| | P-mJOA MDUE subscore | $r < 0$ | $\vert r\vert \geq 0.5$ | -0.28, -0.40 | 12 | +, + | -, - | No |
| | P-mJOA MDLE subscore | $r < 0$ | $\vert r\vert < 0.3$ | -0.42, -0.18 | 12 | +, + | -, + | No |
| | P-mJOA SDUE subscore | $r < 0$ | $\vert r\vert < 0.3$ | -0.28, -0.36 | 12 | +, + | +, - | No |
| | P-mJOA SD subscore | $r < 0$ | $\vert r\vert < 0.3$ | -0.54, -0.44 | 12 | +, + | -, - | No |
| | WHOQOL-Bref EH subscore | $r < 0$ | $\vert r\vert < 0.3$ | +0.21, -0.05 | 12 | +, - | +, + | No |
| | WHOQOL-Bref PH subscore | $r < 0$ | $\vert r\vert < 0.3$ | -0.39, -0.38 | 12 | +, + | -, - | No |
| | WHOQOL-Bref PS subscore | $r < 0$ | $\vert r\vert < 0.3$ | -0.64, -0.43 | 12 | +, + | -, - | No |
| | WHOQOL-Bref SR subscore | $r < 0$ | $\vert r\vert < 0.3$ | -0.15, -0.20 | 12 | +, + | +, + | No |
| Proportion in correspondence: |  |  |  |  |  | 9/9, 8/9 | 4/9, 4/9 | No |
| MoveMed Hold Test | P-mJOA total score | $r > 0$ | $0.3 \leq \vert r\vert (< 0.5)$ | +0.02, +0.55 | 13 | +, + | -, + | No |
| | P-mJOA MDUE subscore | $r > 0$ | $\vert r\vert \geq 0.5$ | +0.14, +0.64 | 13 | +, + | -, + | No |
| | P-mJOA MDLE subscore | $r > 0$ | $\vert r\vert < 0.3$ | +0.13, +0.36 | 13 | +, + | +, - | No |
| | P-mJOA SDUE subscore | $r > 0$ | $\vert r\vert < 0.3$ | +0.21, +0.47 | 13 | +, + | +, - | No |
| | P-mJOA SD subscore | $r > 0$ | $\vert r\vert < 0.3$ | -0.45, +0.14 | 13 | +, + | -, + | No |
| | WHOQOL-Bref EH subscore | $r > 0$ | $\vert r\vert < 0.3$ | +0.14, +0.31 | 21 | +, + | +, - | No |
| | WHOQOL-Bref PH subscore | $r > 0$ | $\vert r\vert < 0.3$ | +0.16, +0.43 | 21 | +, + | +, - | No |
| | WHOQOL-Bref PS subscore | $r > 0$ | $\vert r\vert < 0.3$ | -0.18, +0.02 | 21 | -, + | +, + | No |
| | WHOQOL-Bref SR subscore | $r > 0$ | $\vert r\vert < 0.3$ | +0.31, +0.47 | 21 | +, + | -, - | No |
| Proportion in correspondence: |  |  |  |  |  | 8/9, 9/9 | 5/9, 4/9 | No |
| MoveMed Typing Test | P-mJOA total score | $r > 0$ | $0.3 \leq \vert r\vert (< 0.5)$ | +0.38 | 20 | + | + | No |
| | P-mJOA MDUE subscore | $r > 0$ | $\vert r\vert \geq 0.5$ | +0.37 | 20 | + | - | No |
| | P-mJOA MDLE subscore | $r > 0$ | $\vert r\vert < 0.3$ | +0.21 | 20 | + | + | No |
| | P-mJOA SDUE subscore | $r > 0$ | $\vert r\vert < 0.3$ | +0.32 | 20 | + | – | No |
| | P-mJOA SD subscore | $r > 0$ | $\vert r\vert < 0.3$ | +0.07 | 20 | + | + | No |
| | WHOQOL-Bref EH subscore | $r > 0$ | $\vert r\vert < 0.3$ | +0.08 | 20 | + | + | No |
| | WHOQOL-Bref PH subscore | $r > 0$ | $\vert r\vert < 0.3$ | +0.17 | 20 | + | + | No |
| | WHOQOL-Bref PS subscore | $r > 0$ | $\vert r\vert < 0.3$ | +0.15 | 20 | + | + | No |
| | WHOQOL-Bref SR subscore | $r > 0$ | $\vert r\vert < 0.3$ | +0.10 | 20 | + | + | No |
| Proportion in correspondence: |  |  |  |  |  | 9/9 | 7/9 | No |
| MoveMed Stand and Walk Test | P-mJOA total score | $r > 0$ | $0.3 \leq \vert r\vert (< 0.5)$ | –0.04 | 21 | – | – | No |
| | P-mJOA MDUE subscore | $r > 0$ | $\vert r\vert < 0.3$ | –0.17 | 21 | – | + | No |
| | P-mJOA MDLE subscore | $r > 0$ | $0.3 \leq r (< 0.5)$ | +0.35 | 22 | + | + | No |
| | P-mJOA SDUE subscore | $r > 0$ | $\vert r\vert < 0.3$ | –0.18 | 21 | – | + | No |
| | P-mJOA SD subscore | $r > 0$ | $\vert r\vert < 0.3$ | –0.14 | 21 | – | + | No |
| | WHOQOL-Bref EH subscore | $r > 0$ | $\vert r\vert < 0.3$ | –0.28 | 21 | – | + | No |
| | WHOQOL-Bref PH subscore | $r > 0$ | $\vert r\vert < 0.3$ | –0.12 | 21 | – | + | No |
| | WHOQOL-Bref PS subscore | $r > 0$ | $\vert r\vert < 0.3$ | 0.00 | 21 | ? | + | No |
| | WHOQOL-Bref SR subscore | $r > 0$ | $\vert r\vert < 0.3$ | –0.31 | 21 | – | – | No |
| Proportion in correspondence: |  |  |  |  |  | 1/8 | 7/9 | No |

\*Data reported as single ' $r$ ' values or 'dominant hand, non-dominant hand' $r$ pairs.
†'+' = Sufficient, '–' = Insufficient, '?' = Indeterminate. Data reported as single ratings or 'dominant hand, nondominant hand' rating pairs.
EH: Environmental health, MDLE: Motor dysfunction of the lower extremity, MDUE: Motor dysfunction of the upper extremity, P-mJOA: Patient-derived modified Japanese Orthopaedic Association, PH: Physical health, PS: Psychological health, ROB: Risk of bias, SR: Social relationships, WHOQOL-Bref: World Health Organization quality of life brief version

### Correlations with patient-reported comparators

Spearman rank correlation coefficients are reported in Table 2. As expected, correlations were positive between PerfO’s and PRO’s measuring directly related constructs (e.g., Hold and Typing Test vs. P-mJOA and WHOQOL-Bref) and negative between PerfO’s and PRO’s measuring inversely related constructs (e.g., Fast Tap Test vs. P-mJOA and WHOQOL-Bref). This was most pronounced in the Fast Tap, Hold, and Typing Tests.

Correlation magnitudes were, furthermore, highest between PerfOM’s and PROM’s of neuromuscular function (e.g., Fast Tap Test vs. P-mJOA ≥ 0.3) and lowest between PerfOM’s and PROM’s of quality of life (e.g., Fast Tap Test vs. WHOQOL-Bref < 0.3).

This was also in accordance with expectation, and most pronounced in the Fast Tap, Hold, and Typing Tests.

Correlation magnitudes were notably low (<0.3) in the Stand and Walk Test. This could be due to it being the only multidimensional PerfOM in the battery. Importantly, correlation with the lower limb comparator domain (i.e., the P-mJOA MDLE subscore) was highest, in accordance with expectation.

### Risk of bias assessment

No risk of bias factors from the COSMIN Risk of Bias checklist were recorded. This was equivalent to a ‘very good’ rating for methodological quality (31).

### Overall assessment

Hypotheses and result ratings are also reported in Table 2. These are appraised in writing due to the relatively higher weight of some comparators over others.

Over 70% of correlations for the Fast Tap, Hold, and Typing Tests were in correspondence with hypotheses (Table 2). This provides robust evidence for the validity of these PerfOM’s in the assessment of DCM: particularly due to the relatively higher importance of the correlations to the upper limb comparator (i.e., the P-mJOA MDUE subscore), which were concordant. For the Stand and Walk Test, 50% of the correlations were in correspondence with hypotheses. This also provides evidence for the validity of this PerfOM in the assessment of DCM: particularly due to the relatively higher importance of the correlation to the lower limb comparator (i.e., the P-mJOA MDLE subscore), which was concordant. Taken together, these data provide ‘very good’ quality evidence for the overall validity of the PerfOM’s in the assessment of DCM.

## DISCUSSION

Smartphone applications are increasingly being used to administer clinical outcome measures in medicine. This study used consensus-based standards to assess the validity of an application designed by neurosurgeons and computer scientists from the University of Cambridge. Two lines of evidence were produced. First, a panel of correlations between the application’s tasks and established clinical comparators, and second, a panel of ratings made in accordance with pre-specified hypotheses. The former produced modular evidence of construct validity and the latter a means for its overall appraisal. This type of evidence corresponds to clinical validation under the V3 framework for BioMeTs, and succeeds laboratory-based verification and analytical validation (20).

Construct validity uses comparison to other measures to assess validity. Where comparators take different approaches or contain their own limitations, validity should not be defined by traditional correlation thresholds (35). This is applicable to DCM, where we are trying to improve disease measurement. For example, we recognise the mJOA as a gold standard measure of disease severity, but it measures multiple constructs with limited discrimination, particularly of milder disease. If a new measure had a correlation of 1.0, it would simply mean the instruments are equivalent but consequently very unlikely to be an improvement. For construct validity, exploring expected relationships through hypothesis testing is therefore preferred. As expected, the direction and magnitude of MoveMed’s correlations were most convergent between tasks and questionnaires measuring similar constructs than tasks and questionnaires measuring dissimilar construct (e.g., Fast Tap Test vs. P-mJOA > Fast Tap Test vs. WHOQOL-Bref). This is because neuromuscular tasks should correlate more with neuromuscular constructs than with non-neuromuscular ones (e.g., finger dexterity vs. upper extremity neuromuscular function > finger dexterity vs. quality of life) and because unidimensional tasks should correlate more with other unidimensional measures than with multidimensional ones (e.g., unidimensional vs. unidimensional > unidimensional vs. multidimensional).

To enable performance across correlations to be judged, a proportion of overall hypothesis agreement may be used (32). After rating, >70% and 50% of unidimensional and multidimensional results were respectively deemed sufficient for construct validity in the DCM subgroup. In the absence of risk of bias factors, these data provide ‘very good’ quality evidence for the validity of MoveMed’s tasks in DCM.

The standards adopted by this study have been previously used in the assessment of PerfOM’s by authors of the COSMIN guidance (31). Whilst not originally designed for this purpose, these standards are considered to be a cornerstone in clinimetric validation and, importantly, overlap with industry guidance from the United States Food and Drug Administration (16, 36). This study thus made a point to conduct and report the COSMIN Risk of Bias assessment to aid the reader in their interpretation of the rating panels (Table 2).

Whilst construct validity testing (often criterion validity) is more commonly used by investigators, correlation coefficients require interpretation, as outlined. For similar reasons, the relative performance of instruments should not be judged solely based on the magnitude of correlation coefficients. This is reflected in clinimetric standards who instead recognise <u>content</u> validity as the most important arbitrator of validity and wider performance. Content validity uses stakeholder judgement and feedback to determine validity, and will be further reported on separately for MoveMed, following study completion. When developing and reviewing measurement instruments, understanding clinimetrics is therefore critical.

In this cohort, the impact that DCM would be expected to have on the P-mJOA and WHOQOL-Bref was similarly seen on the Fast Tap, Hold, Typing, and Stand and Walk MoveMed Tests. Correlations with total scores and limb-specific subscores were recorded, in accordance with pre-specified expectations. The most interesting finding was the strong correlation between the P-mJOA MDUE subscore and the Hold Test Stability Score. This is because upper limb stability is not classically thought to be a marker of DCM. The authors attribute this finding to the composite nature of the upper limb stability construct, which includes elements of arm strength, muscle fatigue, and balance. Further studies will follow up with more data on the subject (e.g., content validity). This may very well be an example of a subclinical phenomenon that the human eye cannot catch but that mobile sensors can.

An important strength of this study is its design by individuals with formal training in clinimetrics. This is reflected in the absence of risk of bias factors from the COSMIN checklist in Table 2, and the study’s reporting. There is, unfortunately, a general paucity of well-designed clinimetric studies in the literature (31, 32, 37–39). Use of the COSMIN manual is thus strongly encouraged by the authors. Another strength in this study was the use of PerfOMs that can collect several measurements quickly, ecologically, and longitudinally. This means that the construct should be captured more precisely, more reflective of pathology in the patient’s natural environment, and potentially more responsive to intervention. In the future, these hypotheses will be formally assessed via further clinimetric studies.

Despite its conscientious design, this study has limitations. First, standards for patient-reported methods were adapted to assess performance-based methods. This was done to overcome the absence of standardised criteria in this field and because there is precedent for it in Terwee et al (32) and the COSMIN manual (31). Second, this study reports on 27 individuals (seven months of recruitment). The COSMIN standards are known for being rigorous (and/or stringent) and, ideally, at least 50 participants should be included to earn a modified GRADE score of ‘High’ (31, 32, 37–39). Third, we assumed that the constructs of all WHOQOL-Bref domains would be dissimilar to the PerfOM’s but this may not be the case. The WHOQOL-Bref, ultimately, contains questions on physical activity and the relatedness of this construct to the Fast Tap Test’s and the Typing Test’s may have been observed in Table 2. Fourth, people with a severe form of the disease may have been excluded from enrolment. This would be due to the exclusion of individuals that were unable to stand and walk without the assistance of another person. The potential risks of remote participation in this subset of individuals, however, were deemed to outweigh the benefits by the ethical committee. Further in-person research could address this limitation in the future.

## CONCLUSIONS

The current study provides initial evidence for the validity of the MoveMed PerfOM’s in the context of adults living with DCM in community.

## DATA AVAILABILITY STATEMENT

All data relevant to the study are included in the article.

## ETHICS STATEMENT

This study has been independently assessed and approved by the University of Cambridge (HBREC.2022.13).

## CONTRIBUTORS

BMD and AYT conceived the study and are the guarantors. AYT, BMD, TH, and ZR collected and analysed the data. AYT and BMD wrote the manuscript. IL, KM, ARM, ND, ZG, MK, and MRNK provided critical and independent appraisal of the methods, data, and manuscript. All authors reviewed and critically revised the manuscript prior to submission.

## FUNDING

This study was supported by MoveMed Ltd.

## COMPETING INTERESTS

All authors have completed the ICMJE uniform disclosure form at www.icmje.org/coi_disclosure.pdf and declare: support from MoveMed Ltd for the submitted work; BMD is Chief Executive Officer of MoveMed Ltd; AYT is Chief Scientific Officer of MoveMed Ltd; MRNK is Chief Medical Officer of MoveMed Ltd; MK is Chief Data Officer at MoveMed Ltd; IL has received consultation fees from DePuy Synthes, Royalties, Globus and acted as an advisor to Chiefy Inc; ARM has received research grants from AO Spine; ND is the principal investigator of a prospective DCM study in Canada; no other relationships or activities that could appear to have influenced the submitted work.

## ACKNOWLEDGEMENTS

We thank the ongoing support of all participants and stakeholders, including Myelopathy.org (DCM Charity; www.myelopathy.org).

## Notes

### Funding Statement

This study was funded by MoveMed Ltd.

### Author Declarations

Ethics committee of University of Cambridge gave ethical approval for this work (HBREC.2022.13).

